# Vaccination Status and Its Association with Complications among the Patients Admitted with Measles in the Dedicated Measles Hospitals, Dhaka, Bangladesh

**DOI:** 10.64898/2026.06.24.26356486

**Authors:** Arpita Goutam, Anamul Hasan, Md Abdullah Saeed Khan, Mohammad Ali Hossain Mahid, Sumaya Binte Masud, Habiba Babul, Lamisa Rahman, Amina Siddiqui, Salma Tasnim Luthfa, Samiha Nahar Tuli, Indrani Paul, M Rashidul Bari, Abu Sayeed Md Saleh, Mohammad Delwer Hossain Hawlader

**Affiliations:** Public Health Promotion and Development Society, Crescent Road, Dhaka-1205, Bangladesh; Department of Public Health, North South University, Dhaka 1229, Bangladesh; NSU Global Health Institute (NGHI), North South University, Dhaka 1229, Bangladesh; International Centre for Diarrhoeal Disease Research, Bangladesh (icddr,b), Dhaka-1212, Bangladesh; BRAC James P Grant School of Public Health, BRAC University, Dhaka, 1212, Bangladesh; Division of Clinical and Translational Research, Department of Medicine, McGill University; Directorate General of Health Services (DGHS), Dhaka, 1212, Bangladesh; Mahidol Oxford Tropical Medicine Research Unit, Faculty of Tropical Medicine, Mahidol University, Thailand; Centre for Tropical Medicine and Global Health, University of Oxford, Oxford, UK; World Health Organization, Bangladesh, Dhaka 1212; Eastern Medical College, Kabila, Cumilla, Bangladesh; Ibn Sina Medical College Hospital, Kallyanpur, Dhaka 1216, Bangladesh

**Keywords:** Measles, child health, Vaccination, Complication, Hospitalization

## Abstract

Bangladesh is facing a major resurgence of measles, with more than 60,000 suspected cases and over 600 deaths reported between March and May 2026. Despite the growing outbreak, hospital-based evidence in Bangladesh remains limited regarding measles vaccination status and its association with clinical complications. To address this critical gap, our study aimed to assess the vaccination status and its relationship with the development of clinical complications.

A total of 260 children admitted to the measles ward were enrolled in this cross-sectional study. They were aged 2–72 months, had clinically confirmed measles, and were admitted to four dedicated measles treatment hospitals in Dhaka, Bangladesh, between 15 and 25 April 2026. Data on vaccination status, sociodemographic characteristics, feeding practices, Nutritional status, clinical symptoms, and complications were collected through caregiver interviews and hospital records. Adjusted odds ratios (AOR) with a corresponding 95% confidence interval (CI), and a p-value of <0.05 were considered statistically significant.

Among enrolled children, 74.6% were unvaccinated, 18.8% were partially vaccinated, and only 6.5% were fully vaccinated. In the multivariable model, age below 9 months (aOR 0.077, 95% CI 0.025–0.236,p<0.001) was independently associated with lower odds of vaccination, while household income at or above the median (aOR 3.480, 95% CI 1.493–8.110,p=0.004) was associated with higher odds. Complications developed in 31.1% of cases, with respiratory involvement being most common. Absence of exclusive breastfeeding (aOR 2.336, 95% CI 1.027–5.313,p=0.043) and presenting with exactly three symptoms at admission (aOR 3.106, 95% CI 1.274–7.572,p=0.013) were independently associated with complications. Unvaccinated individuals exhibited markedly elevated odds of complications compared to those who were vaccinated (aOR 5.729,95% CI: 2.363-13.889, p<0.001).

The overwhelming burden of measles in unvaccinated children, shaped by socioeconomic disadvantage and suboptimal feeding practices, underscores the urgent need to restore immunization coverage and strengthen equitable health services in Bangladesh.

## Introduction

Measles, a highly contagious respiratory disease caused by a virus in the paramoxyviridae family, remains a leading vaccine-preventable contributor of childhood morbidity and mortality worldwide, particularly in low- and middle-income countries [1, 2]. The virus is transmitted through respiratory droplets and aerosols with an incubation period of 10-14 days. Clinical manifestations include fever, cough, coryza, conjunctivitis, and Koplik spots, later followed by maculopapular rashes, which may lead to common complications like ear infections, diarrhea as well as severe outcomes encephalitis, blindness, and death, especially among unvaccinated children under five years of age [3]. Despite the availability of safe, effective, and affordable measles-containing vaccines (MCV), the disease has not yet been completely eradicated, and, unfortunately, progress achieved over decades of immunization efforts has been reversed [4].

Globally, measles vaccination averted around 59 million deaths between 2000 and 2024 [5, 6], demonstrating a significant impact on immunization campaigns, but the recent substantial upsurge has become a significant public health threat. Immunization disruptions during the COVID-19 pandemic have exacerbated the global resurgence of measles, healthcare inequities, missed vaccinations and widening immunity gaps among children [7–9]. According to the World Health Organization (WHO) and the U.S. Centers for Disease Control and Prevention (CDC), an estimated 10.3 million people worldwide were infected with measles in 2023, an increase of about 20% compared to 2022 [5, 10]. During the same period, 57 countries reported major measles outbreaks, indicating an increase of about 60% compared to the previous year. In 2024, the first-dose coverage was 84%, while the second-dose coverage was around 74%, both substantially below the 95% threshold to establish herd immunity to halt disease transmission and prevent estimated deaths of 95,000, mostly among unvaccinated or partially vaccinated children under five [5, 11].

The situation in the South Asian region is currently one of the most vulnerable areas for measles transmission due to its dense population, limited health care infrastructure, malnutrition and uneven vaccination coverage [12]. Particularly, countries like Pakistan, Afghanistan, India, and Bangladesh have witnessed repeated measles recrudescence. In many parts of the region, barriers to vaccination, including vaccine hesitancy, lack of awareness, corruption, and sometimes, parental refusal influenced by religious beliefs, significantly increase both susceptibility to infection and disease severity [13–16]. However, vaccination has been shown to play a critical role in reducing disease severity, as vaccinated individuals are less likely to transmit the virus and tend to experience milder clinical outcomes compared to unvaccinated individuals [1, 7].

Bangladesh has achieved considerable progress in reducing measles-related morbidity and mortality through the Expanded Program on Immunization (EPI) and nationwide mass measles-rubella (MR) immunization campaigns [17], with coverage of the first and second doses of measles vaccine reaching 94 and 93%, respectively, in 2016. The benefits of vaccination have been clearly visible over the years, with only 7,000 suspected cases reported nationwide in 2023. However, a recent outbreak causing suspected 60,540 cases and suspected 414 deaths to be reported between 15th March and 22nd May 2026 [18], suggests a plausible lapse in the immunization chain [19, 20]. Amid recent political unrest, measles vaccine coverage declined from over 90% in 2020 to around 57% in 2025 [21], which explains the resurgence. Vaccine stockouts, shortage of health workers, the absence of nationwide immunization campaigns since 2020, and structural inequities such as poverty, low maternal education, higher birth order, and child malnutrition significantly influenced vaccination uptake [19, 22]. Consequently, these outbreaks have overwhelmed healthcare facilities, leading to a concentration of numerous hospitalizations and deaths in dedicated measles centers.

Despite extensive evidence highlighting the role of vaccination in preventing measles and mitigating disease severity [23, 24], there remains limited facility-based research in Bangladesh examining how vaccination status influences the occurrence of complications among hospitalized patients. This gap is particularly significant in outbreak settings, where healthcare facilities concentrate severe cases and may have limited treatment capacity. Therefore, this study aimed to assess the vaccination status and its association with complications among patients with measles who were hospitalized in the dedicated measles hospitals in Bangladesh. The findings are expected to aid in improving clinical management, strengthening immunization strategies, and informing public health interventions aimed at reducing measles-related morbidity and mortality.

## Methodology

### Study design and participants

This hospital-based cross-sectional study was conducted from 15-25 April 2026 among 260 children aged 2 to 72 months who had confirmed measles and were admitted to dedicated treatment facilities in Dhaka, Bangladesh: Dhaka Medical College Hospital, Infectious Disease Hospital, Shaheed Suhrawardy Medical College Hospital, and DNCC dedicated Covid-19 Hospital. Participants were recruited consecutively at the time of admission from the measles wards. Measles cases were confirmed based on a standard clinical case definition, including fever, maculopapular rash, and at least one of cough, coryza, or conjunctivitis, in accordance with WHO guidelines, with laboratory confirmation by measles-specific IgM antibody detection using enzyme-linked immunosorbent assay (ELISA) where available [25]. Data were collected from parents or legal guardians through caregiver interviews and supplemented by hospital records using a structured questionnaire and case record form (CRF). Pediatric patients were eligible if a caregiver or legal guardian was present and willing to provide informed consent for the interview and review of hospital records. Patients were excluded if they were discharged before data collection was completed, if the guardian or care giver declined informed consent, if essential medical documentation was incomplete, if the patient or caregiver refused to continue the interview, or if they had immunodeficiency diseases or were undergoing immunosuppressive therapy.

### Study Procedure

Data were collected using a structured questionnaire and a clinical data extraction checklist derived from hospital case records or documentation. Trained researchers conducted face-to-face interviews with caregivers of children who had been admitted to the measles wards with confirmed measles in the selected dedicated measles treatment hospitals in Bangladesh. The study was explained to the caregivers, and informed consent was obtained. Vaccination cards were reviewed when available. Information on complications and other study variables was extracted from hospital case records and treatment sheets. Data were collected using Kobo Toolbox. Interviewers clarified items as needed to ensure accurate understanding and responses.

### Questionnaire

For data collection, we used a questionnaire to conduct face-to-face interviews that consisted of both closed-ended and semi-structured questions. The questionnaire consisted of five major sections. The first section collected general information, including participant identification number, date of interview, name of measles center/hospital and date of hospital admission.

The second section gathered socio-demographic information of the child and caregiver, including age, sex, religion, place of residence, anthropometric measurements (weight, height, and mid-upper arm circumference), caregiver’s educational status and occupation and monthly household income. The third section focused on vaccination-related information. Data regarding vaccination status were verified by vaccine cards, caregiver recall or hospital records. Information was collected on the child’s receipt of age-appropriate vaccines, measles-containing vaccine first dose (MCV-1), second dose (MCV-2), and 5-year booster dose for age-eligible children. Reasons for non-vaccination and age at vaccination were also documented. The fourth section assessed the nutritional status of the child. Information regarding exclusive breastfeeding, formula feeding, combined feeding practices, nutritional status retrieved from patient records, and vitamin A supplementation after admission was collected.

The final section documented clinical symptoms, disease complications, and patient outcomes. Clinical presentations at admission, including fever, rash, cough, runny nose, and conjunctivitis, were recorded. Information regarding the development of complications, type of complications, and the outcome of illness since admission was also included in the questionnaire. Additionally, respondents were allowed to provide further comments.

The questionnaire was finalized after pretesting to check for the appropriateness and validity of the variables used in the study.

### Study variables and measurements

The primary dependent variable of the study was the complications among hospitalized measles patients. Complications were categorized into respiratory system complications (pneumonia, difficulty breathing, and cough), gastrointestinal complications (diarrhoea, dehydration, and loss of appetite), and auditory complications such as otitis media.

The independent variables included vaccination, nutritional, and feeding-related characteristics. The vaccination status was verified through three sources: vaccination cards (when available), caregiver recall, and hospital records. Based on measles-containing vaccine (MCV) uptake, participants were categorized as unvaccinated (Those who did not get any measles-containing vaccine), partially vaccinated (Those who got only measles-containing vaccine, first dose MCV-1) & fully vaccinated (Those who got both MCV-1 and MCV-2). In logistic regression analysis, we combined people who were partially vaccinated and fully vaccinated into one group called “vaccinated”. People who hadn’t received any dose stayed in the “unvaccinated” group to clearly see its association with complications. Feeding history was categorized into exclusive breastfeeding, formula feeding, breastfeeding with formula feeding & complementary feeding.

Other variables, such as age, have been expressed in months and categorized into three groups: less than 9 months, 9–15 months, and above 15 months. Caregiver occupational status was grouped into blue-collar/informal work (RMG worker, factory worker, street vendor), white-collar/formal employment (private or public service holder), and self-employed/trade (shopkeeper or businessperson). Caregiver educational status was categorized as primary (Class I–V), secondary (Class VI–SSC), and higher secondary (HSC and above). Bangladesh has eight administrative divisions. For analytical convenience, place of residence was categorized into inside the Dhaka division and outside the Dhaka division, the latter comprising the remaining seven divisions. The income group was categorized based on the median value of the monthly household income of 23,000 BDT. Based on this, two groups were created: below median group & at and above median group.

### Statistical Analysis

Data were entered and cleaned in Microsoft Excel 2019 and analyzed using Stata version 15, R version 4, and GraphPad Prism version 9. Descriptive statistics were used to summarize participant characteristics. Categorical variables were presented as frequencies and percentages, while continuous variables were summarized using mean and standard deviation or median and interquartile range, as appropriate. Logistic regression analysis was performed to assess factors associated with vaccination status and complications. Crude odds ratios and adjusted odds ratios with 95% confidence intervals were reported. Variables considered relevant from the literature and those with statistical significance in univariate analysis were included in the multivariable models. A *p*-value of less than 0.05 was considered statistically significant.

### Ethical statement

Ethical approval for the study was obtained from the Institutional Review Board of North South University, Dhaka (IRB No. 2026/OR-NSU/IRB/0407). Informed consent was obtained from the parents or legal guardians of all enrolled children at the measles wards before data collection. Since participation in this study was voluntary, the participants could withdraw from the interview at any moment. Privacy and confidentiality of all collected information were strictly maintained by the researcher, thereby ensuring that all ethical considerations were met and the privacy rights of human subjects were upheld. The ethical theory transcribed in the 1964 Declaration of Helsinki and its subsequent amendments were followed.

## Results

The sociodemographic and economic profile of the measles cases and their caregivers is summarized in **Table 1**. The median age of the children was 10 months (IQR 8.5 months). Almost half (46.1% n=120) were younger than 9 months, 28.5% (n=74) were 9–15 months old, and 25.4% (n=66) were older than 15 months. Males accounted for 54.2% (n=141) of cases. Most caregivers had secondary-level education (66.9%, n=174), and 51.9% (n=135) were housewives. The median monthly household income was BDT 23,000 (IQR 25,500).

**Table 1:**
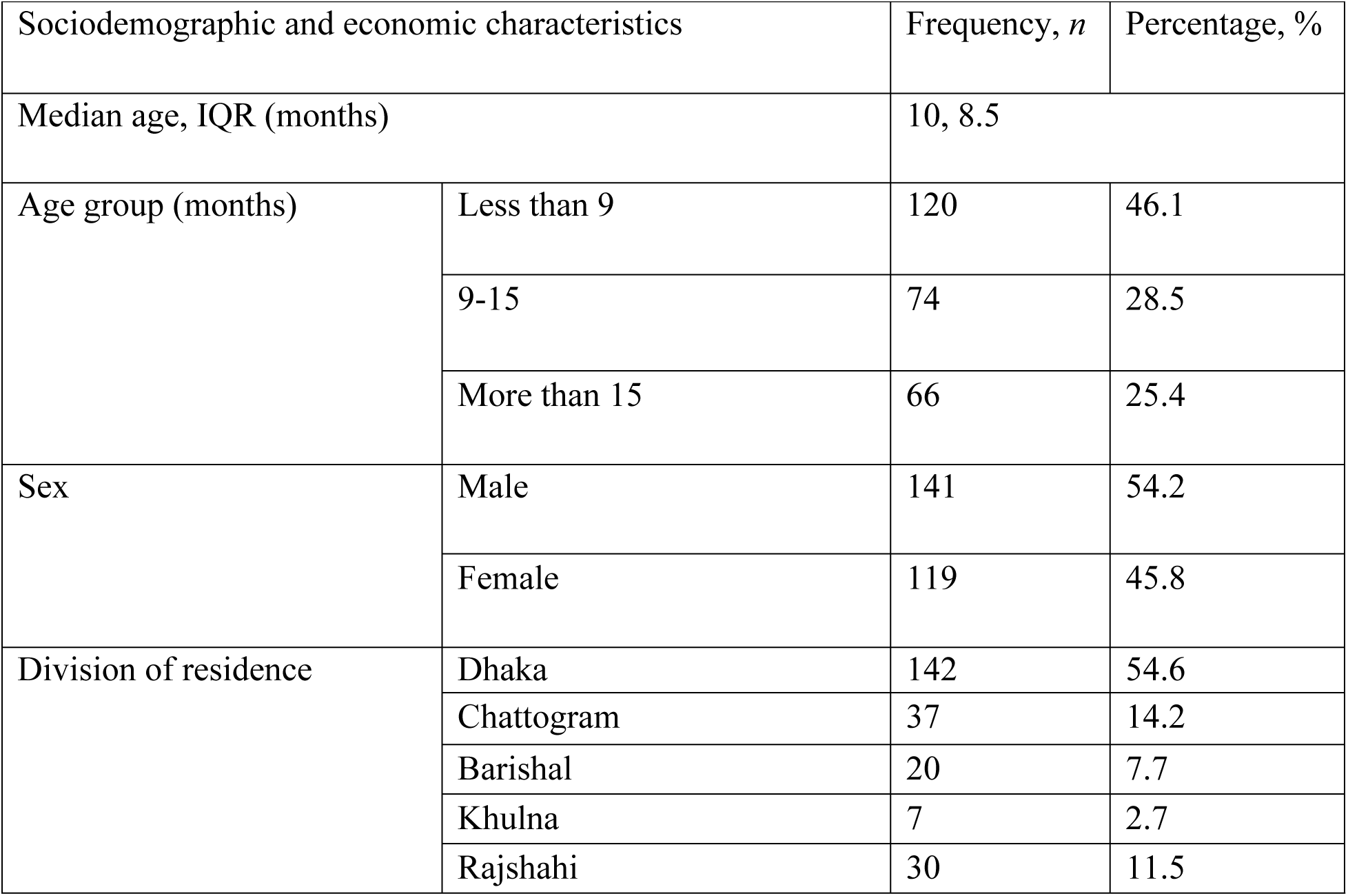
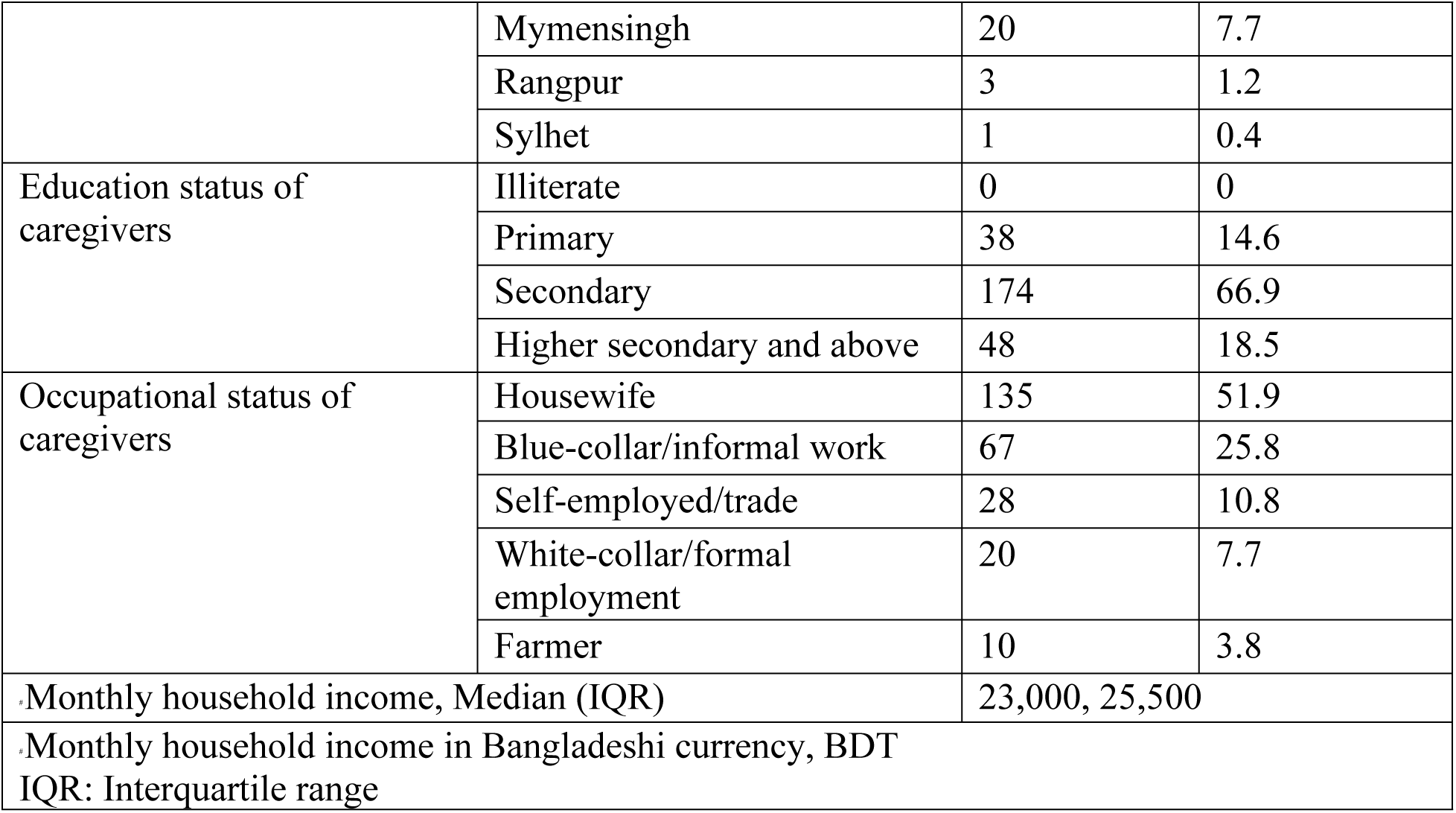
Sociodemographic and economic characteristics of measles cases and caregivers in the study (N = 260)

On the other hand, **Fig 1** shows the district-wise origin of the enrolled measles cases treated in different Dhaka-based government hospitals. A total of 37 out of 64 districts in Bangladesh (57.8%), including Dhaka, were represented among the enrolled patients, indicating that these hospitals served a broad geographic catchment area. The largest numbers of enrolled cases came from Dhaka (n = 58), followed by Narayanganj (n = 22), Cumilla (15), Narsingdi and Tangail (n = 13, each), Gazipur (n = 11), Bhola, Bogura, and Mymensingh (n = 10, each).

**Fig 1:**
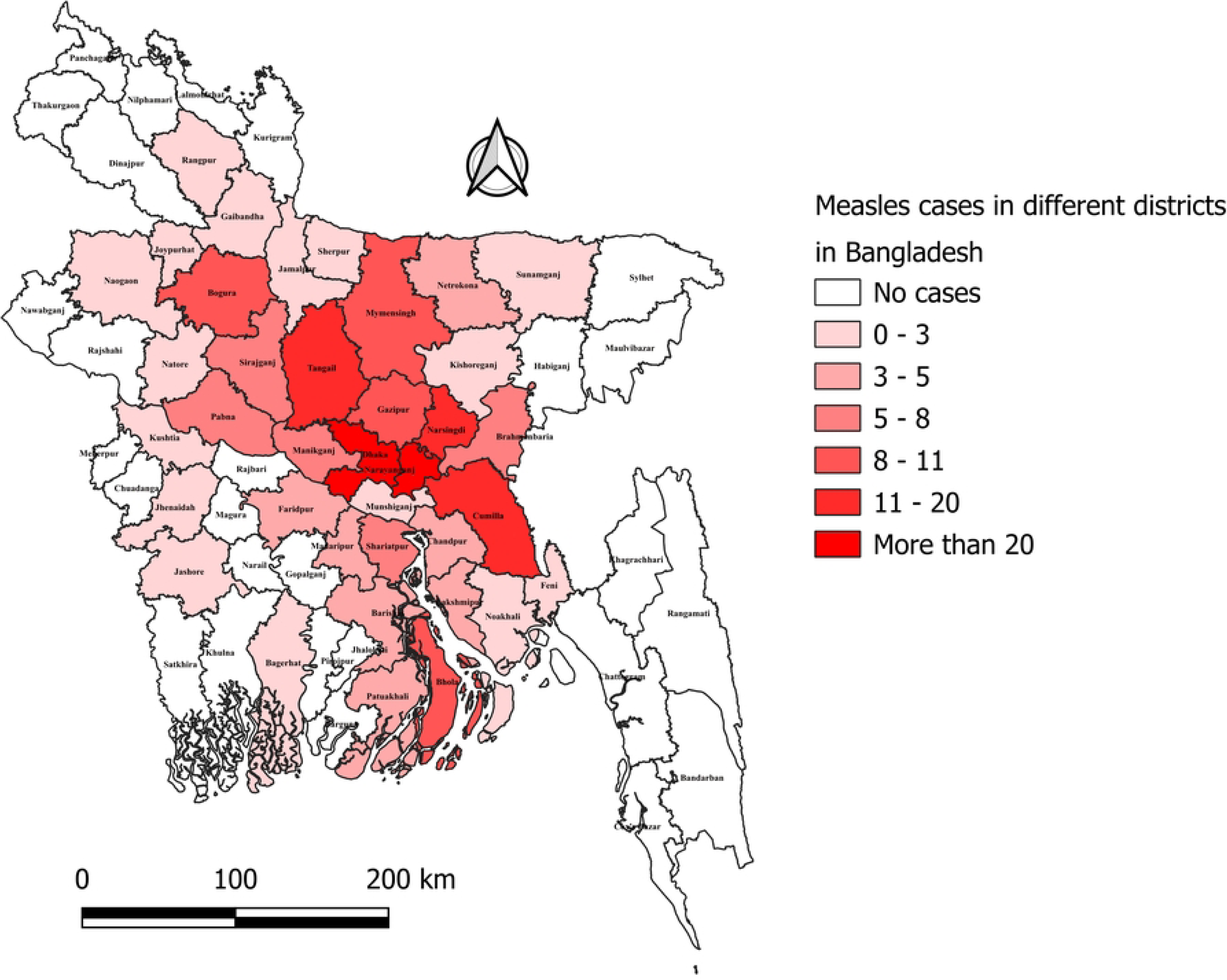
District-wise spatial distribution of enrolled measles cases in the study.

Clinical and nutritional characteristics are presented in **Table 2**. Feeding practices were mixed: 47.3% (n=123) received both exclusive breastfeeding and formula feeding, 24.6% (n=64) were exclusively breastfed, 14.2% (n=37) were on complementary feeding, and 13.9% (n=36) were on formula feeding only. Nutritional status was normal in 89.6% (n=233) of cases. Vitamin A supplementation after admission was documented for 85.4% (n=222) of children. All participants presented with fever and rash. Cough was reported in 74.6% (n=194), runny nose in 28.1% (n=73), and conjunctivitis in 18.5% (n=48). Complications developed in 31.1% (n=81) of cases. Among those with complications, respiratory-system involvement was most common (66.7%, n=54), followed by gastrointestinal (27.1%, n=22), auditory (3.7%, n=3), and combined respiratory plus gastrointestinal (2.5%, n=2).

**Table 2:**
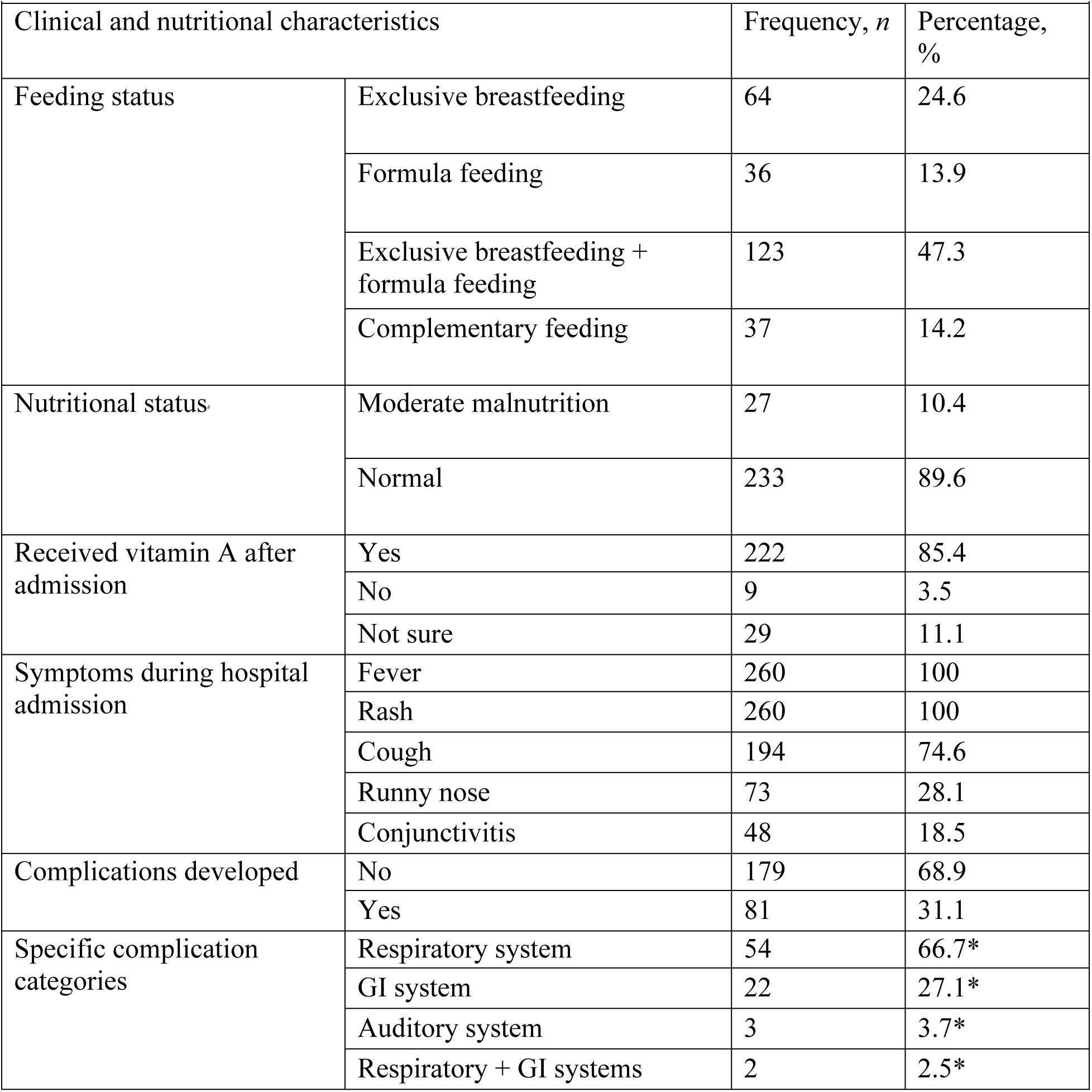
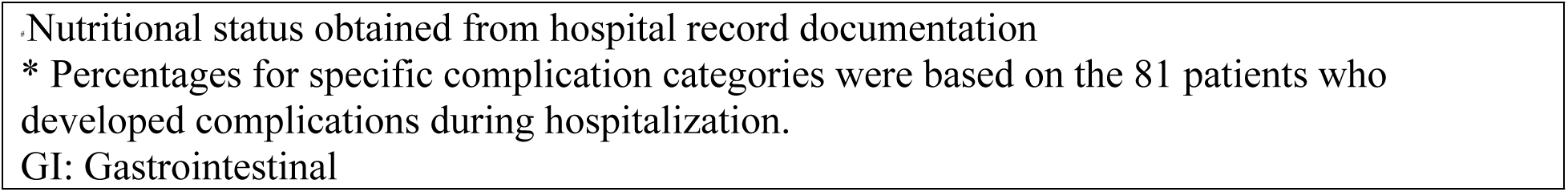
Clinical and nutritional characteristics of measles cases in the study (*N* = 260)

The proportions of different vaccination statuses (unvaccinated, partially vaccinated, and fully vaccinated) among study participants are shown in **Fig 2** (pie chart). Among all the study participants, 194 children (74.6%) were unvaccinated, 49 (18.8%) were partially vaccinated, and only 17 (6.5%) were fully vaccinated against measles.

**Fig 2:**
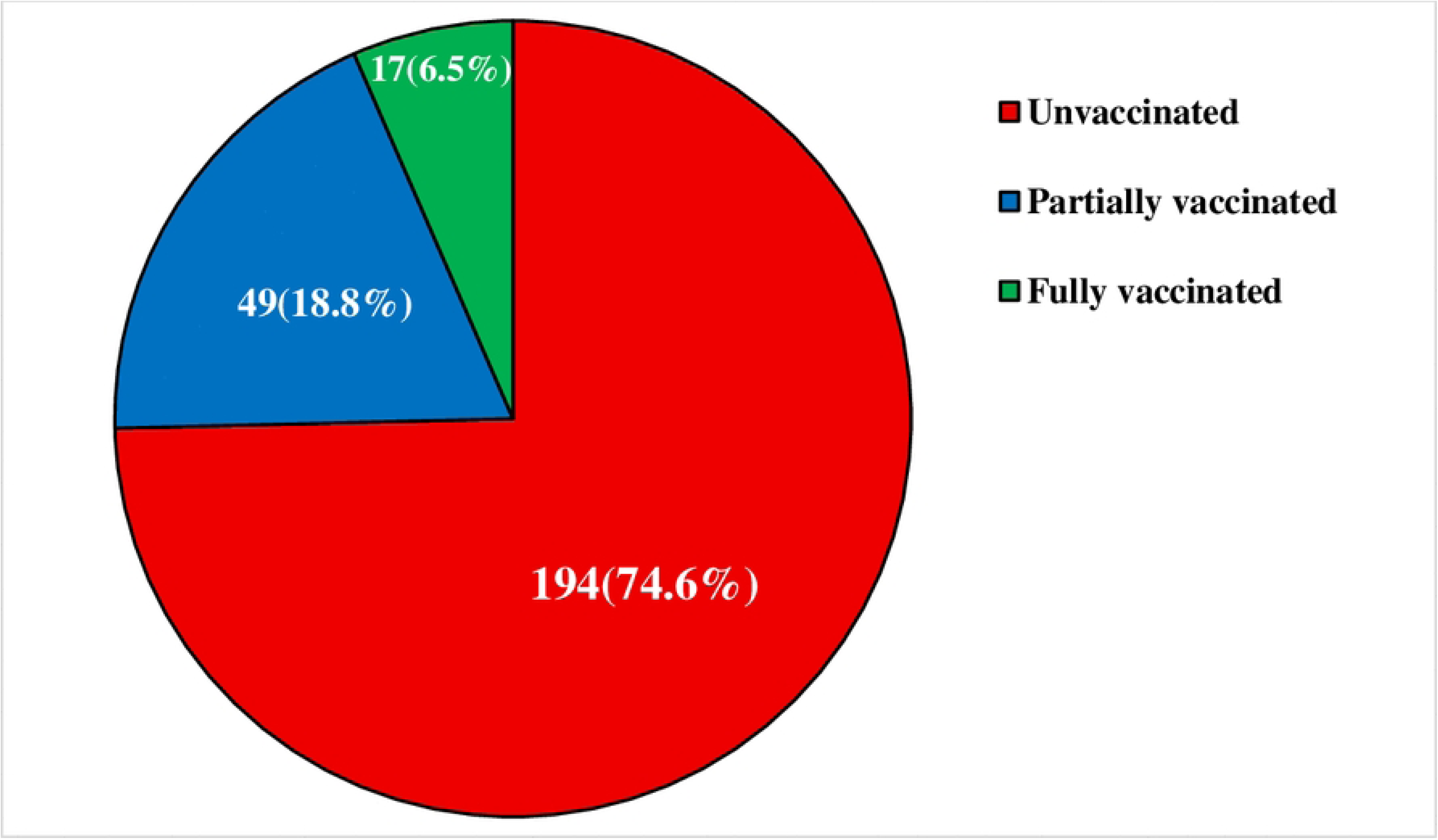
Proportion of vaccination status among the study participants.

Univariate and multivariable logistic regression results for factors associated with vaccination status are shown in **Table 3**. In the adjusted model, children younger than 9 months had significantly lower odds of vaccination compared with the 9–15 months reference group (aOR 0.077, 95% CI 0.025–0.236, p < 0.001). Household income at or above the median was independently associated with higher odds of vaccination (aOR 3.480, 95% CI 1.493–8.110, p=0.004). After adjustment, gender, division of residence, exclusive breastfeeding history, and caregiver education and occupation were not significantly associated with vaccination status.

**Table 3:** Univariate and multivariable logistic regression of factors associated with vaccination status among the study participants.

| Vaccination status | cOR (95% CI) | *p*-value | aOR (95% CI) | *p*-value |
| --- | --- | --- | --- | --- |
| **Gender** |  |  |  |  |
| Male (RC) |  |  |  |  |
| Female | 0.768 (0.436 – 1.352) | 0.360 | 0.515 (0.236 – 1.123) | 0.095 |
| **Age group (in months)** |  |  |  |  |
| 9-15 (RC) |  |  |  |  |
| <9 | 0.103 (0.037 – 0.286) | <0.001 | 0.077 (0.025 – 0.236) | <0.001 |
| >15 | 3.414 (1.696 – 6.872) | 0.001 | 2.404 (0.799 – 7.234) | 0.119 |
| **Division of residence** |  |  |  |  |
| Inside Dhaka (RC) |  |  |  |  |
| Outside Dhaka | 2.492 (1.403 – 4.426) | 0.002 | 1.609 (0.730 – 3.547) | 0.238 |
| **Income group** |  |  |  |  |
| Below median (RC) |  |  |  |  |
| At or above the median | 4.011 (2.113 - 7.617) | <0.001 | 3.480 (1.493 - 8.110) | 0.004 |
| **Exclusive breastfeeding history** |  |  |  |  |
| Exclusive breastfed (RC) |  |  |  |  |
| Not exclusively breastfed | 4.635 (2.545 - 8.441) | <0.001 | 1.420 (0.468 - 4.304) | 0.536 |
| **Caregiver educational status** |  |  |  |  |
| **Primary (RC)** |  |  |  |  |
| Higher education | 0.744 (0.262 - 2.107) | 0.577 | 0.956 (0.202 - 4.530) | 0.955 |
| Secondary | 1.228 (0.541 - 2.783) | 0.623 | 1.244 (0.398 - 3.892) | 0.707 |
| **Caregiver occupational status** |  |  |  |  |
| Housewife (RC) |  |  |  |  |
| Blue-collar/informal work | 0.546 (0.274 - 1.089) | 0.086 | 0.757 (0.286 - 2.003) | 0.575 |
| Farmer | 0.230 (.028 - 1.871) | 0.169 | 0.206 (0.012 - 3.674) | 0.282 |
| Self-employed/trade | 0.248 (0.071 - 0.867) | 0.029 | 0.459 (0.094 - 2.234) | 0.335 |
| White-collar/formal employment | 0.517 (0.163 - 1.638) | 0.262 | 2.079 (0.339 - 12.758) | 0.429 |
| cOR: crude Odds Ratio, aOR: adjusted Odds Ratio, CI: Confidence Interval, RC: Reference Category |  |  |  |  |

**Fig 3** illustrates the distribution of symptom counts (categorized as <3 symptoms, exactly 3 symptoms, or >3 symptoms) at hospital admission, stratified by vaccination status (unvaccinated, partially vaccinated, and fully vaccinated). Unvaccinated patients had the highest number of symptoms, with 89 cases reporting exactly 3 symptoms and 77 cases having more than 3 symptoms. In contrast, partially vaccinated and fully vaccinated patients experienced fewer symptoms overall, with only 27 and 8 cases reporting exactly 3 symptoms, and just 9 and 4 cases having more than 3 symptoms, respectively. The distribution of symptom burden differed significantly by vaccination status(p=0.025). Across all symptom categories, unvaccinated children consistently represented the largest proportion of cases, suggesting a possible relationship between lack of vaccination and more severe clinical presentation among hospitalized measles patients

**Fig 3:**
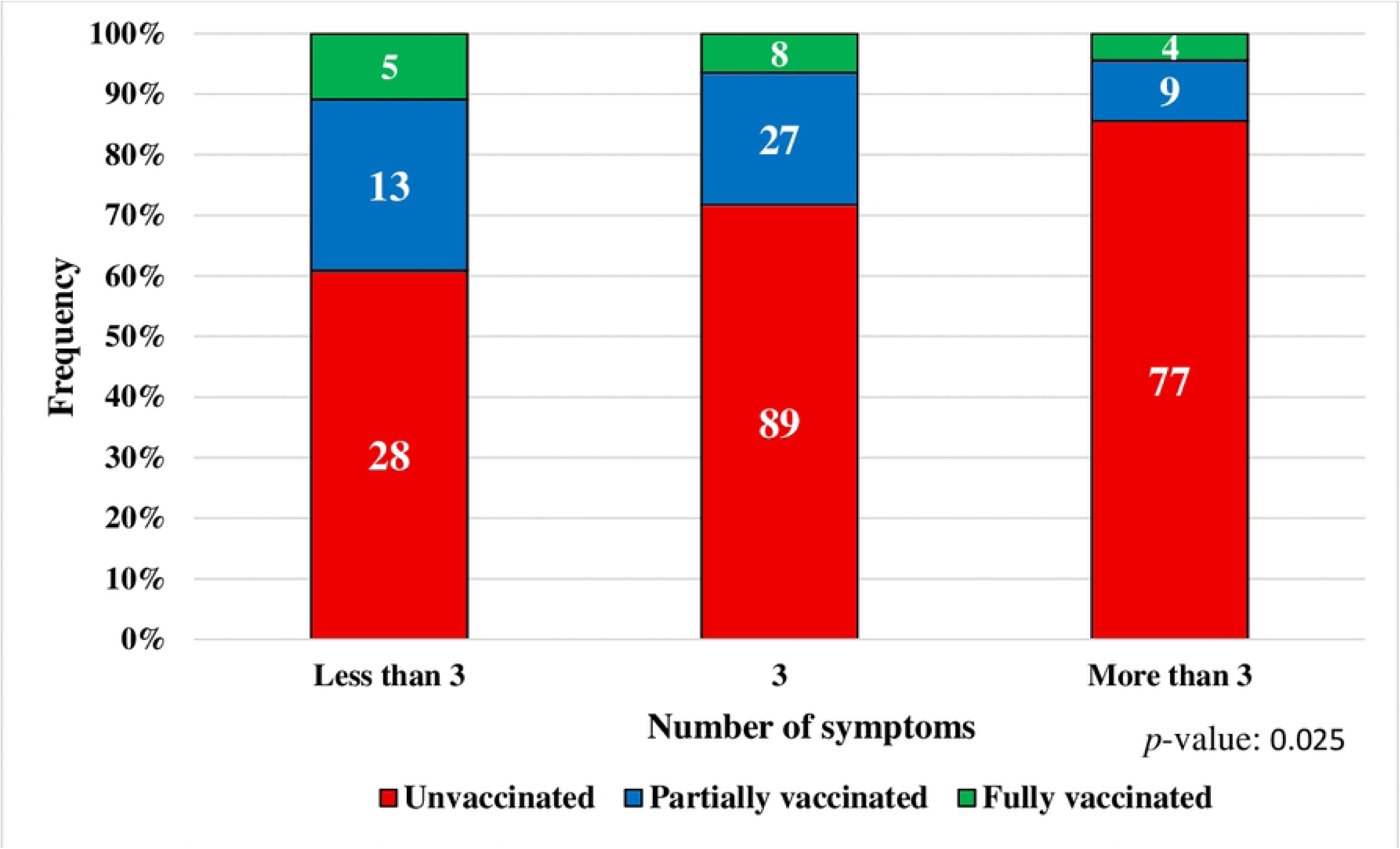
Number of symptoms during hospital admission by vaccination status among measles cases.

**Table 4** presents the univariate and multivariable logistic regression analyses of factors associated with the development of any complications during hospitalization. Not being exclusively breastfed remained independently associated with higher odds of complications (aOR 2.336, 95% CI 1.027–5.313, p=0.043). Children presenting with exactly three symptoms at admission also had significantly increased odds of complications compared with those having fewer than three symptoms (aOR 3.106, 95% CI 1.274–7.572, p=0.013). Unvaccinated individuals had significantly higher odds of developing complications compared to vaccinated individuals (aOR 5.729,95% CI: 2.363-13.889, p<0.001).

**Table 4:** Univariate and multivariable logistic regression of factors associated with complications developed among the study participants.

|  |  |  |  |  |
| --- | --- | --- | --- | --- |
|  | cOR (95% CI) | *p*-value | aOR (95% CI) | *p*-value |
| **Gender** |  |  |  |  |
| Male (RC) |  |  |  |  |
| Female | 0.995 (0.587 - 1.685) | 0.984 | 0.836 (0.469 - 1.491) | 0.544 |
| **Age group (in months)** |  |  |  |  |
| 9-15 (RC) |  |  |  |  |
| <9 | 0.824 (0.439 - 1.544) | 0.545 | 0.578 (0.290 - 1.153) | 0.120 |
| >15 | 1.114 (0.552 - 2.249) | 0.763 | 1.082 (0.399 - 2.929) | 0.877 |
| **Division of residence** |  |  |  |  |
| Inside Dhaka (RC) |  |  |  |  |
| Outside Dhaka | 0.880 (0.518 - 1.493) | 0.636 | 0.809 (0.451 - 1.450) | 0.476 |
| **Exclusive breastfeeding history** |  |  |  |  |
| Exclusive breastfed (RC) |  |  |  |  |
| Not exclusive breastfed | 1.711 (0.970 - 3.018) | 0.064 | 2.336 (1.027 - 5.313) | 0.043 |
| **Total number of symptoms during admission** |  |  |  |  |
| Less than 3 (RC) |  |  |  |  |
| 3 | 3.103 (1.335 - 7.211) | 0.008 | 3.106 (1.274 – 7.572) | 0.013 |
| More than 3 | 1.727 (0.706 - 4.224) | 0.231 | 1.382 (0.535 – 3.570) | 0.504 |
| **Vaccination status** |  |  |  |  |
| Vaccinated* (RC) |  |  |  |  |
| Unvaccinated | 2.823 (1.389 – 5.744) | 0.004 | 5.729 (2.363 – 13.889) | <0.001 |
| * "Vaccinated" covers both partially and fully vaccinated cases. |  |  |  |  |
| cOR: crude Odds Ratio, aOR: adjusted Odds Ratio, CI: Confidence Interval, RC: Reference Category |  |  |  |  |

## Discussion

In this study we evaluated the vaccination status of children admitted to dedicated measles treatment facilities in Dhaka, Bangladesh and its association with clinical complications. The results show a substantial immunisation gap: approximately three-quarters of hospitalised children were unvaccinated (74.6%) and only 6.5% were fully vaccinated. Household income above the median was the sole independent predictor of vaccination status in the adjusted model. Nearly half of all cases (46.1%) were infants younger than nine months, the minimum age for measles-containing vaccine (MCV1) under the national immunisation schedule, and were therefore ineligible for vaccination at the time of illness. In the multivariable model for complications, vaccination status, non-exclusive breastfeeding, and a higher symptom burden at admission were independent predictors, with vaccination status showing the strongest association.

The high proportion of unvaccinated children among hospitalised measles cases is consistent with findings from hospital-based studies in comparable low- and middle-income countries and reflects the well-established epidemiological link between lack of vaccination and severe measles requiring hospitalization [24, 26]. This finding must be interpreted carefully. The Expanded Programme on Immunisation (EPI) in Bangladesh has achieved substantial and well-documented gains in measles-containing vaccine coverage over recent decades [27], and the present study was not designed to evaluate overall national programme performance. Recent analyses signal concerning declines in routine immunisation coverage, with the 2026 measles resurgence interpreted by some as evidence that immunisation gains in Bangladesh remain fragile [19, 20]. The resurgence has been linked to vaccine stockouts, EPI procurement disruptions following the political transition of 2024, and periods of frontline health worker industrial action, all of which reduced effective coverage across successive birth cohorts [28, 29]. Against this backdrop, the concentration of unvaccinated cases in tertiary referral hospitals points to residual pockets of vulnerability capable of sustaining localised transmission even in a setting of high aggregate national coverage [20].

Household income at or above the median was independently associated with a more than threefold increase in the odds of vaccination (aOR 3.480; 95% CI 1.493 to 8.110; p=0.004). This finding is consistent with a broad body of evidence from comparable countries demonstrating that lower socioeconomic position reduces access to preventive health services, including vaccines, through multiple pathways such as reduced mobility, lower health-system contact, and competing livelihood priorities [30–33]. After adjustment for income, neither caregiver education, occupational status, nor geographic residence retained statistical significance in the model. The absence of an independent effect of education and occupation after adjustment for income suggests that material resources, rather than knowledge or awareness, are the primary structural pathway through which socioeconomic conditions shape immunisation behaviour in this population. Similarly, the crude association between residence outside Dhaka and higher vaccination odds was attenuated to non-significance after adjustment, indicating that this geographic pattern was attributable to differences in the socioeconomic mix of patients rather than geography per se [34].

A particularly important contextual factor is that nearly half of all enrolled cases (46.1%; n=120) were infants younger than nine months, an age group not yet eligible for MCV1 under the Bangladesh national schedule. This group is biologically dependent on maternally transferred antibodies, which wane progressively during the first months of life, creating a window of susceptibility before the first scheduled vaccine dose can be administered [35, 36]. The concentration of cases in this pre-vaccination age window, consistent with observations from other South Asian countries [37], underlines that strategies to reduce measles morbidity cannot rely on vaccination alone and must incorporate complementary interventions to protect this highly vulnerable infant cohort. These findings link directly to the income-related immunisation gap described above: addressing socioeconomic barriers to vaccination would reduce the burden in children who are eligible but unvaccinated, while separate strategies are needed for those who are pre-vaccination by age.

One of the strongest modifiable risk factors for complications in the adjusted model was non-receipt of exclusive breastfeeding (aOR 2.336; 95% CI 1.027 to 5.313; p=0.043). Respiratory complications were the most common type, affecting 66.7% of children who developed any complication, in keeping with international evidence identifying pneumonia as the leading cause of measles-attributable mortality in young children in low- and middle-income country settings [37]. The protective association of exclusive breastfeeding against respiratory infections in Bangladeshi children has been documented in prior hospital-based work from this setting [38]. The biological basis is well established: breast milk provides secretory immunoglobulin A, lactoferrin, and other bioactive components that support mucosal immunity and limit secondary bacterial infection in infants [39]. Although the cross-sectional design of the present study precludes causal inference, the direction and magnitude of the association are nonetheless consistent with this immunological evidence. The high proportion of cases aged under nine months reinforces the salience of exclusive breastfeeding as a preventive strategy in the very age group that is not yet eligible for vaccination and is therefore most reliant on passive and nutritional protection.

Vaccination status emerged as an independent and statistically significant predictor of complications in the multivariable model. Unvaccinated children had markedly higher odds of developing complications than those who had received one or two doses of measles-containing vaccine (aOR 5.729; 95% CI 2.363 to 13.889; p < 0.001). This is in line with the well-established protective effect of measles-containing vaccines against disease severity [26, 40, 41]. Two doses confer robust humoral and cellular immunity that limit viral replication and reduce the risk of secondary complications [40, 41]. A single dose appears sufficient to prime the immune system, supporting a more effective secondary response on natural exposure to wild-type virus, which may explain why even partial vaccination conferred meaningful protection in this cohort [26]. Comparable dose-dependent reductions in complications have been reported in hospital-based studies from the United States [42], Pakistan [43], and Dhaka itself [44]. The strength and precision of this association identify vaccination status as the most consequential modifiable determinant of complication risk in this study and reinforce the case for prioritizing catch-up immunisation in the current outbreak response.

Presenting with exactly three symptoms at admission (fever, rash, and one additional symptom) was independently associated with higher odds of complications compared with children presenting with fewer than three symptoms (aOR 3.106; 95% CI 1.274 to 7.572; p=0.013). The number of symptoms at admission may therefore serve as a practical, low-cost triage indicator to identify children at elevated risk of clinical deterioration, complementing other clinical and nutritional assessments at the time of admission [45].

The concentration of hospitalised measles cases among unvaccinated children from lower-income households points to a clear and actionable target for the ongoing 2026 outbreak response in Bangladesh, reinforced by the strong and significant association between vaccination status and complications identified in this study. Reducing socioeconomic barriers to vaccination through targeted outreach campaigns, mobile immunisation services in low-income and peri-urban areas, and integration with existing social protection programmes could have a substantial impact on measles-related hospitalisations and complications. Evidence from Bangladesh supports integrated demand-generation strategies involving community health workers and linkage with safety-net programmes [31, 33]. Beyond demand-side barriers, the resurgence also reflects structural governance failures in EPI supply chain management and programme continuity that require urgent attention to prevent recurrence [28, 29]. The need to strengthen completion of the two-dose measles schedule (MCV2 at 15 months) remains important. Immediate priorities in the context of the current outbreak should include strengthening reminder and recall systems, following up defaulters, and planning catch-up campaigns. Additionally, for the large pre-vaccination age group (<9 months), complementary interventions should continue to promote exclusive breastfeeding, protect maternal measles immunity through antenatal care and consider post-exposure prophylaxis or earlier vaccination strategies in the context of active outbreaks as per WHO guidance. The strong association between non-exclusive breastfeeding and complications supports the inclusion of infant feeding history in routine clinical triage of admitted measles patients. Routine vitamin A supplementation and enhanced pneumonia surveillance should remain core components of inpatient measles management [33]. These measures are consistent with the Directorate General of Health Services’ response to the 2026 resurgence and can help safeguard routine immunisation coverage against future disruptions.

### Strengths

This study was conducted among confirmed measles cases from dedicated measles hospitals that serve as major referral centres for severe disease in Bangladesh, ensuring a well-defined and clinically homogeneous study population. Recruitment across multiple administrative divisions strengthens the representativeness of findings within the measles patient population. Combining partially and fully vaccinated children into a single comparator category provided sufficient statistical power to detect a robust association between vaccination status and complications. The dual analytical approach, using multivariable logistic regression to address both study objectives, provides an internally consistent evidence base. Restriction to confirmed measles cases enhances the reliability of the clinical and epidemiological findings.

### Limitations

This hospital-based study of severe measles cases has limited generalisability to the wider population of measles patients, most of whom are managed in community and primary care settings. Complication rates in this cohort are likely higher than in the general measles case population. Vaccination status was ascertained primarily from caregiver recall and vaccination cards, which may result in misclassification. Residual confounding from unmeasured variables such as household sanitation and prior health-seeking behavior may remain after multivariable adjustment. Causal inference is not appropriate given the observational, cross-sectional design.

## Conclusion

A critical immunization gap, reflected by the large number of unvaccinated and partially vaccinated children among hospitalized measles cases, drove the 2026 measles outbreak in Bangladesh. Socioeconomic vulnerability and suboptimal infant feeding practices were independently associated with both lower vaccination coverage and higher risk of complications. These findings suggest that measles severity in this setting is influenced not only by immunization status, but also by broader social and nutritional disadvantages. Improving measles vaccine coverage, particularly among economically disadvantaged populations, remains essential to reducing ongoing transmission in Bangladesh. At the same time, promoting exclusive breastfeeding and ensuring adequate clinical care for hospitalized children may help reduce complications and lessen the impact of future outbreaks.

## Data Availability

The complete metadata set is available upon request from the Chair, North South University Institutional Review Board/ Ethics Review Committee (contact via Dr.Dipak Kumar Mitra, Email address).

## Acknowledgements

We sincerely thank the caregivers of the children who had just been hospitalized at the measles wards during data collection. Your contributions made this research possible.

